# Income-related inequalities and inequities in access to inpatient healthcare in rural Nigeria

**DOI:** 10.64898/2026.02.12.26346153

**Authors:** A.M Yaqoob, A.A. Salisu, Obumneke Ezie

**Affiliations:** Department of Economics & Development Studies, Prime University Abuja, P. M.B 10, Kuje Area Council, FCT-Abuja, Nigeria; Center for Econometrics and Applied Research (CEAR), Ibadan, Oyo State Nigeria; Department of Economics, Bingham University, Karu, Nasarawa State, Nigeria

**Keywords:** Inequalities, inpatient healthcare, public, private, and Concentration Index (CI)

## Abstract

Healthcare inequality remains a major challenge for health systems globally. While previous studies have examined inequalities in specific healthcare services or sub-domains of care, evidence on disparities in access across formal inpatient healthcare settings in Nigeria remains limited. This study examines income-related inequalities and inequities in access to inpatient healthcare among rural households in Nigeria. The study used cross-sectional data from 624 rural households collected in 2022 as part of a human capital research project commissioned by the African Economic Research Consortium (AERC). Information on demographic characteristics, household income, consumption expenditure, health conditions, region of residence, and access to inpatient care was analyzed. Income-related inequalities and need-adjusted inequities were assessed using the Concentration Index (CI) and the Horizontal Inequity (HI) index. Overall, 81.4%, 49.9%, and 18.4% of respondents reported access to inpatient care at public primary, secondary, and tertiary health facilities, respectively, while 32.7% accessed inpatient care at private facilities. Access to public primary and secondary inpatient care decreased with household wealth, whereas access to public tertiary and private inpatient care increased with wealth. Significant pro-poor inequalities and inequities were observed in access to public primary (CI = −0.1054; HI = −0.0374) and secondary (CI = −0.1063; HI = −0.0377) inpatient care. In contrast, access to tertiary (CI = 0.2382; HI = 0.3660) and private (CI = 0.1502; HI = 0.2180) inpatient care exhibited significant pro-rich inequalities and inequities. Decomposition analysis indicated that non-need factors—particularly household economic status and region of residence—were the largest contributors to inequalities in access across all inpatient care types.

**Conclusion:** Inequalities in access to inpatient healthcare were driven mainly by economic status and region of residence.

## 1 Introduction

One of the major challenges facing the global healthcare system today is inequity in access to health service. The policy goal of public health authorities in most countries is to achieve equity in access to health service in order to improve people’s health and overcome inequalities irrespective of socio-economic circumstances [1, 2]. Besides, the right of all people to access quality healthcare service when needed without financial hardship is emphasized in the principle of Universal Health Coverage (UHC) and this right is recognized in the 1978 Alma-Ata declaration of health for all [3, 4]. The Sustainable Development Goal, SDG3 also focused on equitable access to quality health service [5]. However, inequality in healthcare exists when individuals or population groups with comparable health needs are unequally treated arising from differences in socio-economic conditions including education, employment status, income level, gender and ethnicity [6, 3].

Reports show that the disparities in access to health service are disproportionate and widespread across the population groups, race, ethnicity, regions and nations with a greater concern in low-and medium-income countries [7, 8]. In developing countries including Nigeria, wealthier individuals enjoy greater access to quality health service than their poorer counterparts particularly the rural dwellers and people of low socio-economic status [9, 10]. Consequently, these disparities undermine the health status of the affected population leaving the country at large with poor health outcomes.

Reports show that Nigeria is one of the countries with a high burden of communicable and non-communicable diseases (NCDs) [11]. For communicable disease-related deaths such as maternal deaths, the lifetime risk for a Nigerian woman in her reproductive years is 1 in 22, while the average lifetime risk for women in high– and low-income countries are 1 in 4900 and 1 in 180 respectively [12,13]. This enormous difference suggests one of the largest inequalities of any public health statistics [14]. The risk of children dying before the age of one is higher particularly among rural children than their urban counterparts [15,16]. This is because rural dwellers travel long distance to access health service, and are faced with deprivations that hinder their means of transportation, inadequate health resources and personnel, unprofessional conduct of health providers and lower levels of education [16]. For variations in geographic area of residence, notable significant differences in health indices are also documented. For instance, the maternal mortality rates for the Northeast and North-West geopolitical zones in Nigeria are 10 and 4 times respectively higher than those in the South-west [16, 17]. For Non-Communicable Disease (NCDs) – related deaths, Nigeria is one of the countries in sub-Saharan Africa with a high prevalence of NCD deaths accounting for about 30% of all deaths [18]). At 22%, the risk of premature death from cardiovascular diseases, cancers, respiratory diseases, and diabetes among Nigerians aged 30 to 69-years is unacceptably high [18]. The disability-adjusted life years (DALYs) lost to NCDs in general increased by about 21.3% between 2010 and 2019 versus 6.5% for infectious diseases during the same period [18] suggesting that Nigeria is undergoing an epidemiological transition with a rising burden of NCDs. Most of these deaths are attributable to major risk factors such as aging, unhealthy diet, overweight/obesity, hypertension, diabetes mellitus, atherosclerosis and hyperlipidemis and are preventable if the affected individuals have access to important prevention and screening tools [19]. In many low-and lower-middle-income countries healthcare financing relies heavily on out-of-pocket (OOP) expenditure [20]. In Nigeria where majority of the poor live in rural area and earn less than $1.90 a day, out-of-pocket (OOP) payment for healthcare is an inefficient system and limits the poor from accessing quality health service forcing majority of the people into poverty [21, 22].

As part of the efforts to achieve inclusive healthcare financing, Nigeria operates a three-tier of healthcare system with each administered at the local, state and federal level interacting through a referral system [23]. The three-tier system signifies increasing levels of specialization, complex healthcare delivery, and technology advancement, with primary care being the least complex and tertiary care being the most extensive. Primary care serves as the gate keeper providing much of the care, typically a patient’s first interaction with the healthcare system. Secondary healthcare is delivered through local or regional hospitals equipped with appropriate technology. Tertiary care, the highest level of healthcare is provided in institutions such as teaching hospitals and specialized units dedicated to particular populations—including women, children, and individuals with mental health conditions—or to advanced forms of specialized treatment [24]. In spite of the efforts to ensure unrestricted access to healthcare and promote health equity, these efforts did not yield the desired results as Nigeria ranked 142 out of 195 countries in health system performance [25, 26].

Past studies on inequalities in healthcare utilization are concentrated in South Asia and Latin America. They found inequalities in health service utilization driven by differences in socio-economic factors. For example, Xu *et al.* [27] use post healthcare system reform longitudinal data to find pro-rich inequality in the utilization of healthcare service. They also found the need factors to contribute more than 50% to inequality in health services among rural residents, while the socio-economic status, education level and work status make the largest contribution of all the non-need factors. Ramirez *et al.* [28] found the wealth quintile and the type of health insurance to make the largest contribution to inequality in the use of preventive health services. Using data from rural and urban China, Guo *et al.* [29] find income, chronic disease and old age to significantly contribute to inequality in outpatient and inpatient healthcare utilization.

Available studies [30, 31, 16, 32] in Nigeria focused on child and maternal healthcare, while inequalities in healthcare utilization across the care settings in Nigeria are rarely studied. It may be noted that, while efforts towards equitable access to maternal and child healthcare is plausible for addressing the underlying factors responsible for high maternal and child mortality, such efforts provide limited information for policy makers seeking to close the inequality gaps across the entire healthcare system in Nigeria. The focus of this study is to investigate income-related inequalities and inequities in access to inpatient care among households in riral Nigeria.

The rest of the paper is organized as follows: Section 2 discusses sample, sample size and sampling procedure. Section 3 presents the main results, while discussion of the results are presented in Section 4. Section 5 dwells on the conclusion and policy recommendation.

## 2.0 Materials and Methods

### 2.1 Sample, sample size and sampling procedure

This study was conducted in rural Nigeria using a five-stage sampling procedure to obtain a sample for the study. In the first stage, three out of the six geopolitical zones including the Southeast, Southwest and Northeast were randomly selected. The second stage involved a random selection of two states from each of the selected geopolitical zones making a total of six states selected. The selected states comprise of Imo, Ebonyi, Taraba, Bornu, Oyo and Ekiti states. In the third stage, two rural Local Government Areas (LGAs) were selected randomly from each of the selected states making a total of twelve rural LGAs. The fourth stage involved a random selection of two wards from each of the rural LGAs making a total of twenty-four wards selected. In the last stage, twenty-six farming households were randomly selected from each of the selected wards making a total of six hundred and twenty-four (624). The sample size was computed using the International Fund for Agricultural Development (IFAD) protocol given as:

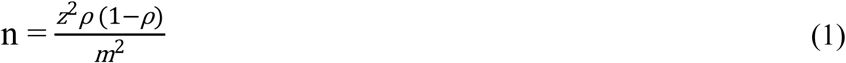

where n = sample size, z = the confidence level at 98% (2.33), *p* = estimated percentage of rural households sampled (65%) and m = the margin of error (4.5% or 0.045). Allowance for design effect and contingency were built into the final sample size (624). The difference between the calculated sample size and the final sample size accounted for design defect and contingency allowance. However, only five hundred and five (505) responses were effectively used for the analysis.

### 2.2 Sources and type of data

The data for the study were sourced from the national rural household survey supported by African Economic Research Consortium (AERC) in conjunction with Bill and Melinda Gates Foundation (BMGF) as part of the commissioned studies for human capital research project in sub-Saharan Africa. The survey, designed to collect data from a sample of rural households in Nigeria, was conducted in six states between July and October 2022. The instrument used for data collection was a semi-structured questionnaire with oral interview. The questionnaire consists of six sections: demographic and socio-economic characteristics (Section A) of the household head; type of illness, inpatient visit to health facilities, and out-of-pocket expenditure on health (Section B), subjective evaluation of healthcare service provider visited (Section C), Household expenditure (Section D) and qualitative responses on households’ assets, housing and dwelling condition (Section E).

### 2.3 Description of variables

#### 2.3.1 Outcome variable

The outcome variable of interest is access to healthcare measured on a binary scale. Access is defined in this study as the opportunity to use and actual utilization of healthcare [33, 3]. Data for various types of healthcare utilization are based on inpatient care. Inpatient care was defined as any use of inpatient health service in the last three months prior the survey period irrespective of the length of hospital stay. The type of healthcare included: (i) public and private healthcare providers. Public healthcare providers are formal healthcare facilities owned by the government providing in-patient and out-patient preventive, curative and restorative health services. They are categorized into the following sub-categories namely primary (dispensaries/maternity centers), secondary (state/general hospital) and tertiary health facilities (teaching/specialist hospital) [34]. Private healthcare are formal health facilities owned by individuals or organization providing in-patient and outpatient preventive, curative and restorative health services to the patients and recognized by the country’s regulatory and legal framework [35].

Access to primary healthcare is a binary variable, taking a value of 1 if the household head or any member of the household had in the past three months prior the survey utilized the service of primary healthcare often referred to as dispensaries/maternity centers and 0 otherwise. Access to secondary healthcare is a binary variable, taking a value of 1 if the household head or any member of the household utilized the service of state or general hospital and 0 otherwise. Access to tertiary healthcare is a binary variable, taking a value of 1 if the household head or any member of the household utilized the service of public teaching or specialist hospital and 0 otherwise. Access to a private health facility is a binary variable taking a value of 1 if household head or any member of the household had in the last three months prior the survey utilized the service of private healthcare and 0 otherwise.

#### 2.3.2 Need Variables

Need variables refer to the health condition as may be perceived or evaluated by individual or household necessitating a visit and use of medical services [36]. We use age, sex and self-reported health condition of the respondent taking a value of 1 if the household head or any member of the household has a chronic health condition (defined as any of the illnesses captured in the study except skin problem), 0 otherwise. These included reproductive illness, polio, asthma, hepatitis, eye/ear infection, acute headache, hypertension, stroke, arthritis, diabetes, skin problem and malaria. Age is a categorical variable taking a value of 1 if the age of the household head is below 30 years, 2 if aged 30-59 years and 3 if aged above 60 years. Sex is a binary variable taking a value of 1 if the household head is male and 0 otherwise.

#### 2.3.3 Predisposing factors (Non-need variables)

Other control variables considered as non-need variables in the study include educational level, marital status, employment type, region of residence and economic status. Educational level is a categorical variable taking a value of 1 if the respondent has at most primary education, 2 if he completed secondary education and 3 if the respondent completed tertiary level of education. Marital status is a binary variable taking a value of 1 if the respondent is married and 0 otherwise. Employment type is a binary variable taking a value of 1 if the primary occupation of the respondent is farming and 0 otherwise. Region of residence is a binary variable taking a value of 1 if the respondent resided in the north east, 2 if resided in the south east and 3 if resided in the southwest. However, for ease of analysis, using the appropriate sample weight, this variable was re-coded into binary variable for statistical analysis taking a value of 1 if the respondent resided in the north and 0 otherwise. We used per capita total household’s expenditure as a measure of household’s economic status classified into five quintile. This is because self-reported income particularly among rural households in Africa and other developing countries are often unreliable due to under-reporting and also prone to seasonal fluctuation.

### 2.4 Statistical Analysis

Following Shen *et al*. [37], concentration index and inequality decomposition were used to achieve the study objectives. The standard measure of the degree of income-related inequality in a given outcome of interest is the Concentration Index (CI). Concentration index (CI) is a relative measure of inequality, defined as twice the covariance between y and fractional rank of a unit in an economic advantage or income distribution r, Cov(y, r), weighted by μ, the mean of y. The value of the CI ranges between –1 and +1. A negative (positive) value indicates that the utilization of a specific healthcare is concentrated among the poor (wealth). A CI equal to zero implies equitable access to healthcare among the various socio-economic groups (Wagstaff *et al.,* 1991). The general model for CI is given as follow:

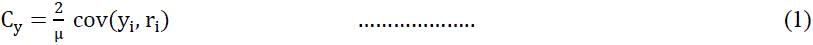

Where C_y_ is the concentration index for healthcare variable y_i_ μ is its mean and r_i_ is the fractional rank of *i^th^* individual unit in income distribution (using per capita consumption expenditure as proxy). Further, a measure of horizontal inequity (HI) developed by Wagstaff *et al.* [38] and Van Doorslaer *et al.* [39] was used to account for the impact of individual needs for the use of health services. HI is defined as the difference between observed health care utilization and need-predicted healthcare use taking into account the fact that individuals have different healthcare needs and that these differences ought to translate into different needs for the use of healthcare. Once health care needs are standardized across individuals, the remaining utilization could be considered to be inequitable.

### 2.5 Decomposition of the Concentration Index

The Concentration Index (CI) was decomposed to determine the relative contribution of each covariate as well as the contribution of unexplained residual component to inequalities in healthcare variable [40, 41]. The decomposition of the CI is performed using a two-part model to identify factors associated with inequality. Binary regression was used in the first part of the Two-part model (TPM) and for the second part, a generalized linear model [42] was estimated. Since the outcome variables are all measured on a binary scale, a linear approximation of a logit model was used to estimate the partial effects:

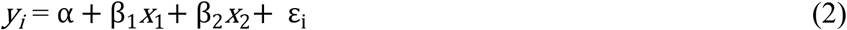

Where α is the intercept, *x*_1_ to *x*_2_ denote the vectors of need and non-need variables (predisposing factors), β_1_and β_2_are the associated parameters and ε_i_is the error term. Therefore, *C_y_*can be written as follow:

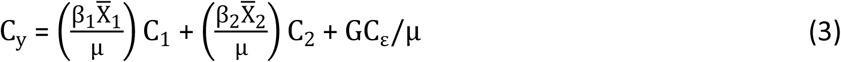

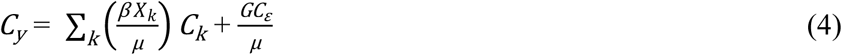

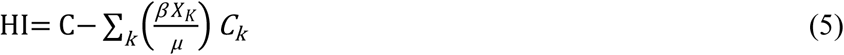

Where 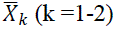 is the mean of each socio-economic factor in a vector, *C_k_* is the CI for *x*_k_ and 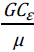 is the generalized CI for the error term. The generalized CI for the error term captures the inequality in access to healthcare that cannot be explained by observed differences between the poor and the rich. We used STATA 17 for the analysis.

### 2.6 Ethical approval and informed consent

Ethical approval for this study was obtained from the National Health Research Ethics Committee (NHREC) prior to data collection. All study procedures were conducted in accordance with relevant ethical guidelines and regulations. Participants were informed about the objectives of the study, and individual written informed consent was obtained. Confidentiality and anonymity of the information provided were assured, and the data were used solely for research purpose.

## 3.0 Results

### 3.1 Descriptive statistics and socio-economic differences in accessing healthcare

The results as presented in Table 1 show that majority of the respondents were male (89.7), married (88.7%), aged 30-59 years (70.9%) with a mean age of 50.18 ± 11.74. Majority of the respondents had at most primary level of education (37.8%). Most of the respondents engaged primarily in farming (51.3%) and had chronic health condition (79.8%). About 81.4% reported access to primary healthcare, 49.9% for secondary healthcare; 18.4% for tertiary healthcare, while 32.7% reported access to private health facilitates [Figure 1].

**Fig. 1:**
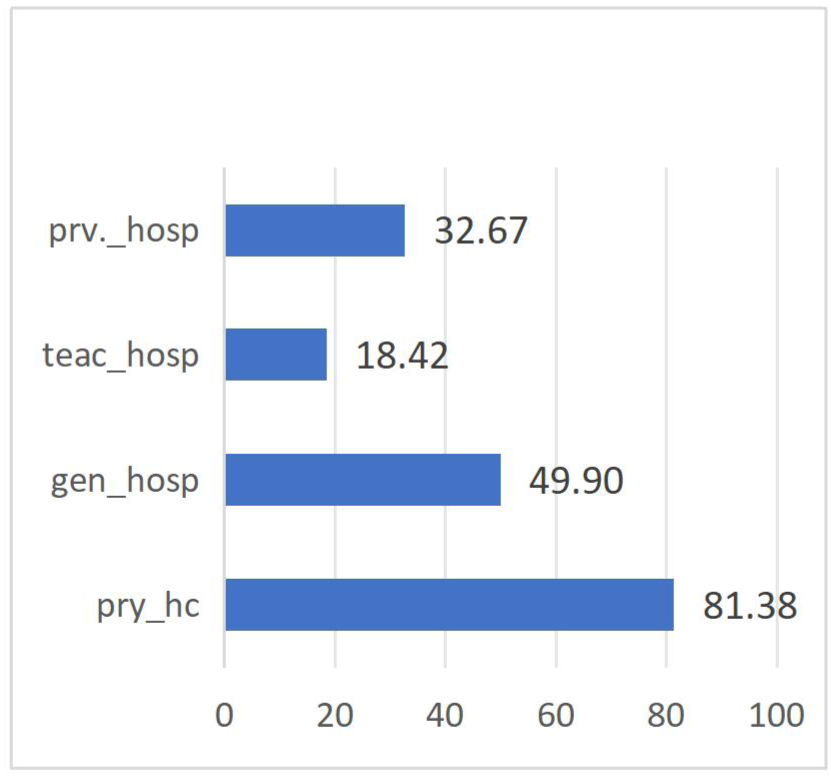
Access to various types of Healthcare. Pry_hc: primary healthcare; gen_hosp: general hospital (secondary); teac_hosp: teaching hospital (tertiary); prv_hosp: private hospital (general healthcare)

**Table 1:**
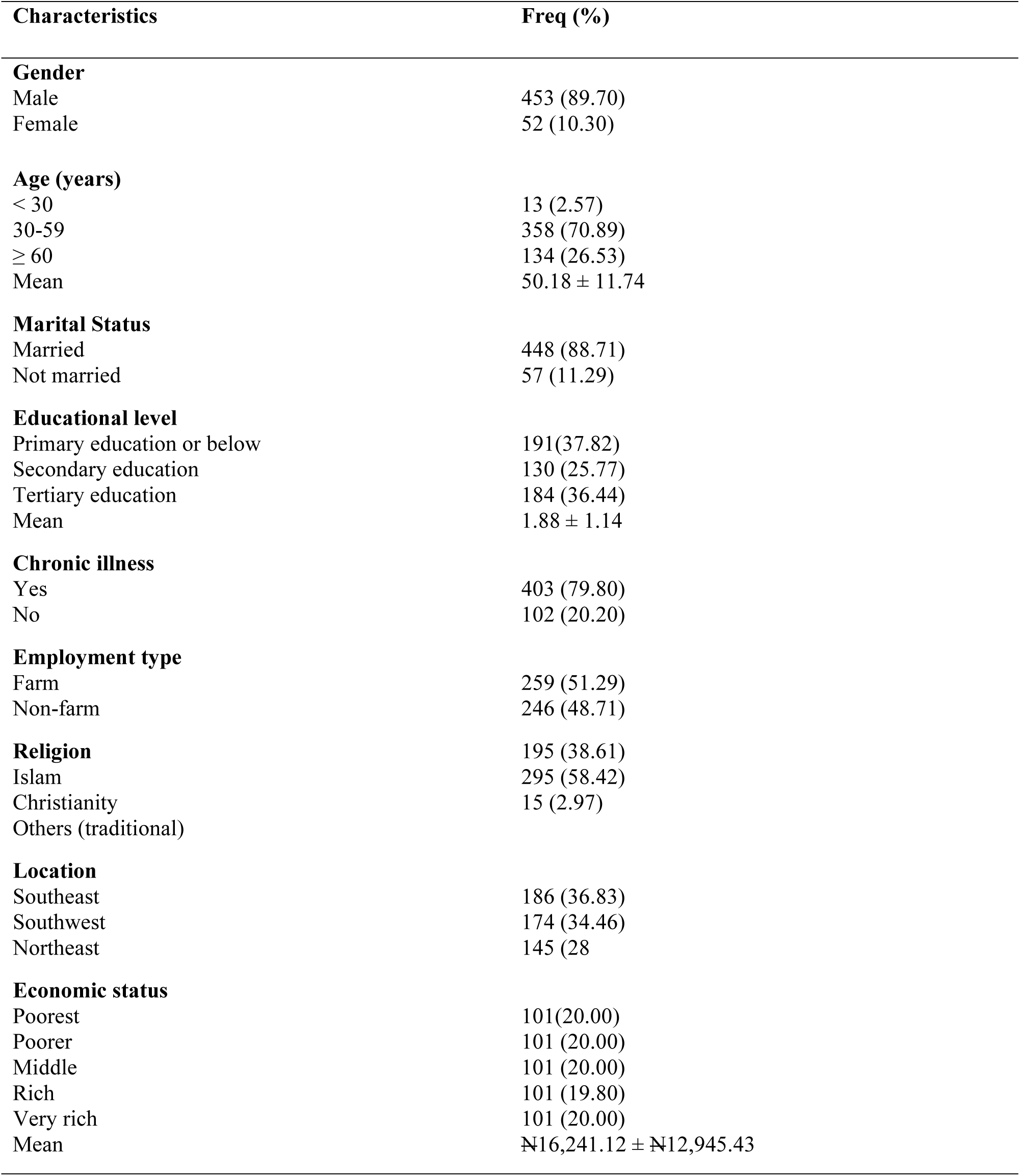
Descriptive statistics of selected variables.

Across the various types of healthcare, access was higher among the male [Public: PHC-91.0%; SHC-92.5%; THC-94.6%; Private: GHC-86.7%], compared to the female [Public:PHC-9.0%; SHC-7.54%; THC-5.38; Private: GHC-13.3%] (Figure 2). Respondents aged 30-59 years (Public: PHC-72.0%; SHC-76.2%; THC-68.8%; Private: GHC-72.7%) and those with completed tertiary educational were found to have the most access to all types of healthcare, while the least access was found among people with less than 30 years and those with at most primary educational attainment (Figure 3-4). The results also show geographical differences in access to healthcare as respondents from the southwest region had the most access to all type of public healthcare, while their counterparts in the south-eastern region had the most access to private facilities only. Overall, north-east was the most-disadvantaged region regardless of the type of healthcare (Figure 5). We found a strong positive correlation between access to healthcare and economic status. For public primary and secondary healthcare, access is decreasing with wealth, while access to tertiary [public] and private general healthcare is increasing with wealth (Figure 6).

**Fig 2:**
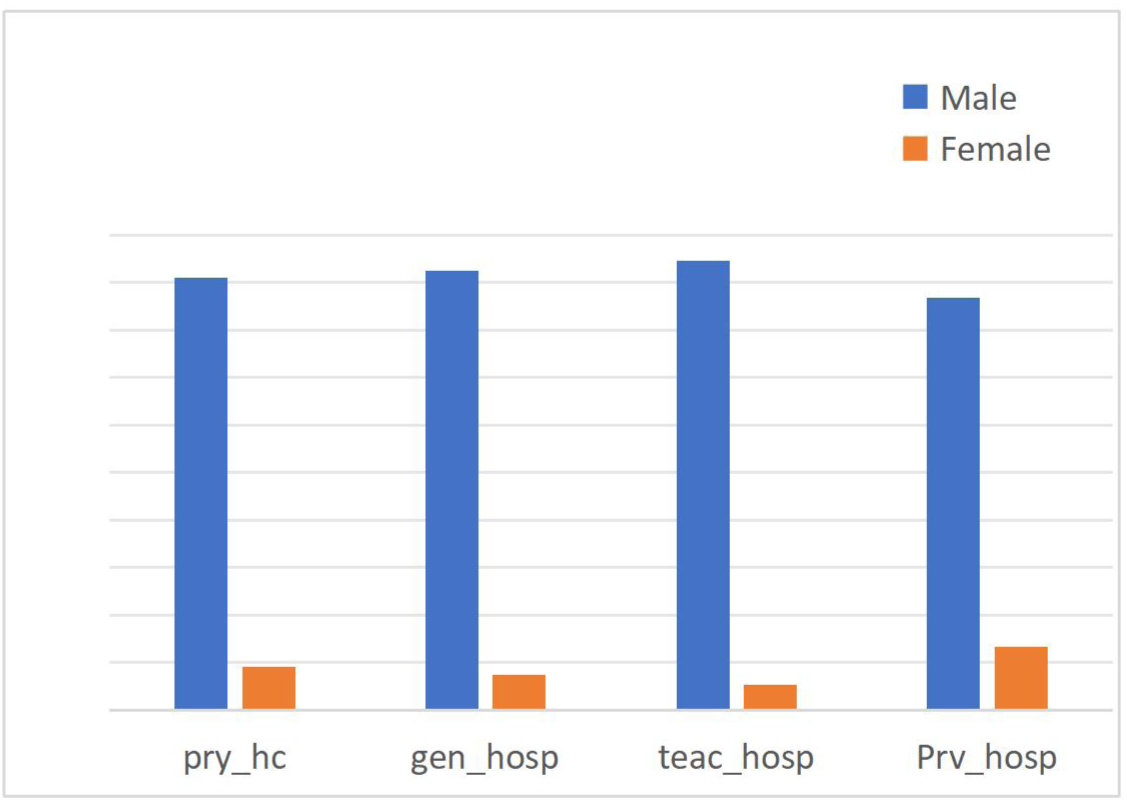
Access to various types of healthcare by gender. Pry_hc: primary healthcare; gen_hosp: general hospital (secondary); teac_hosp: teaching hospital (tertiary); prv_hosp: private hospital (general healthcare)

**Fig. 3:**
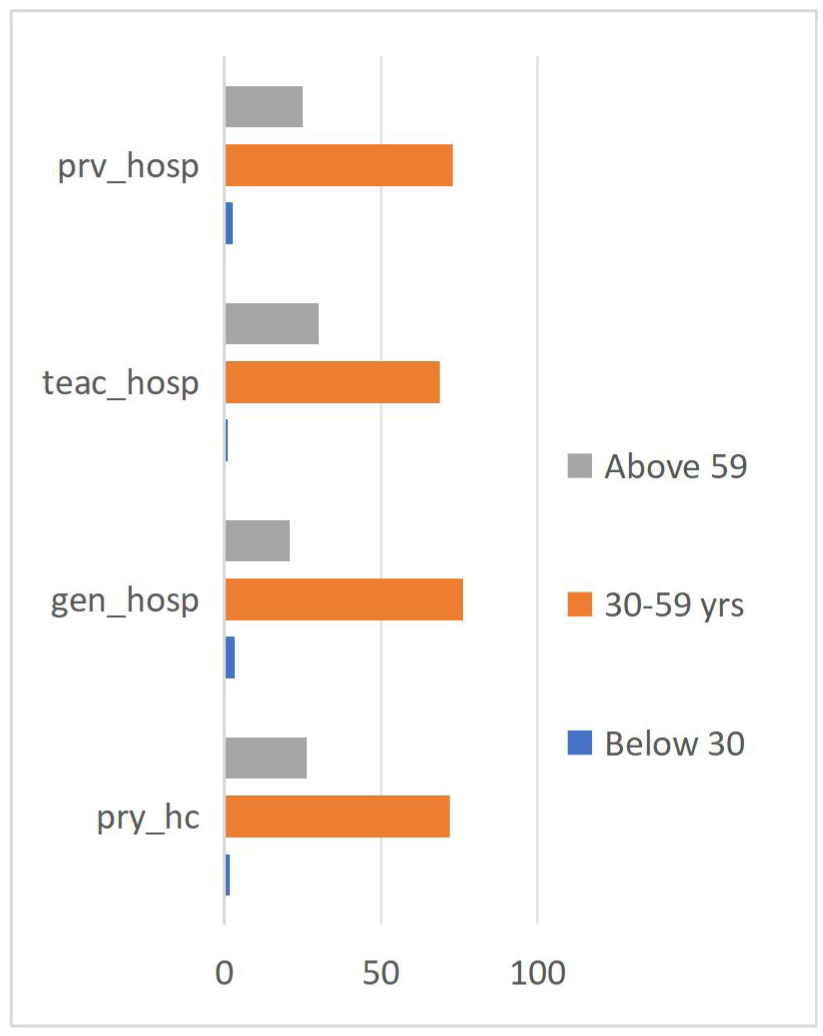
Access to healthcare by age category.

**Fig. 4:**
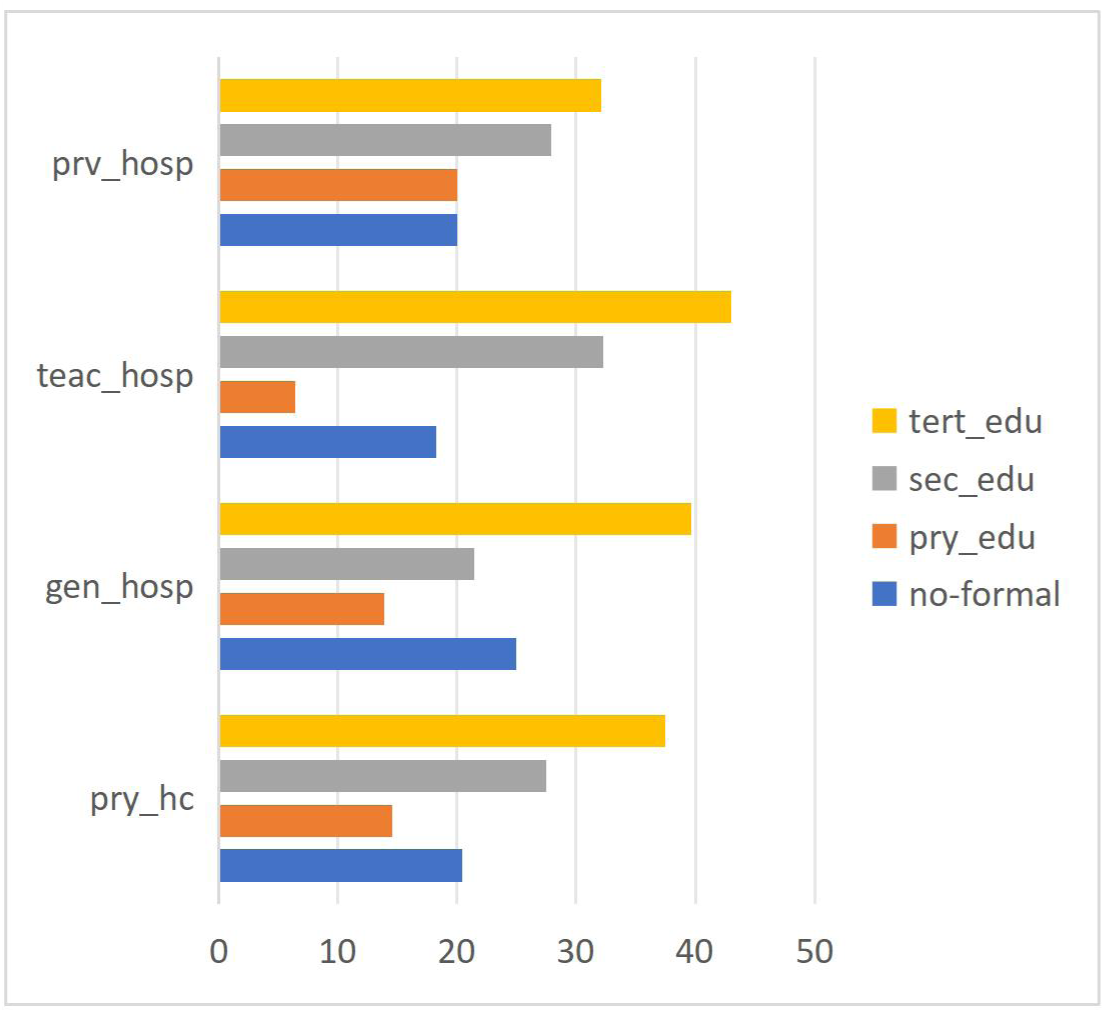
Access to healthcare by educational level.

**Fig. 5:**
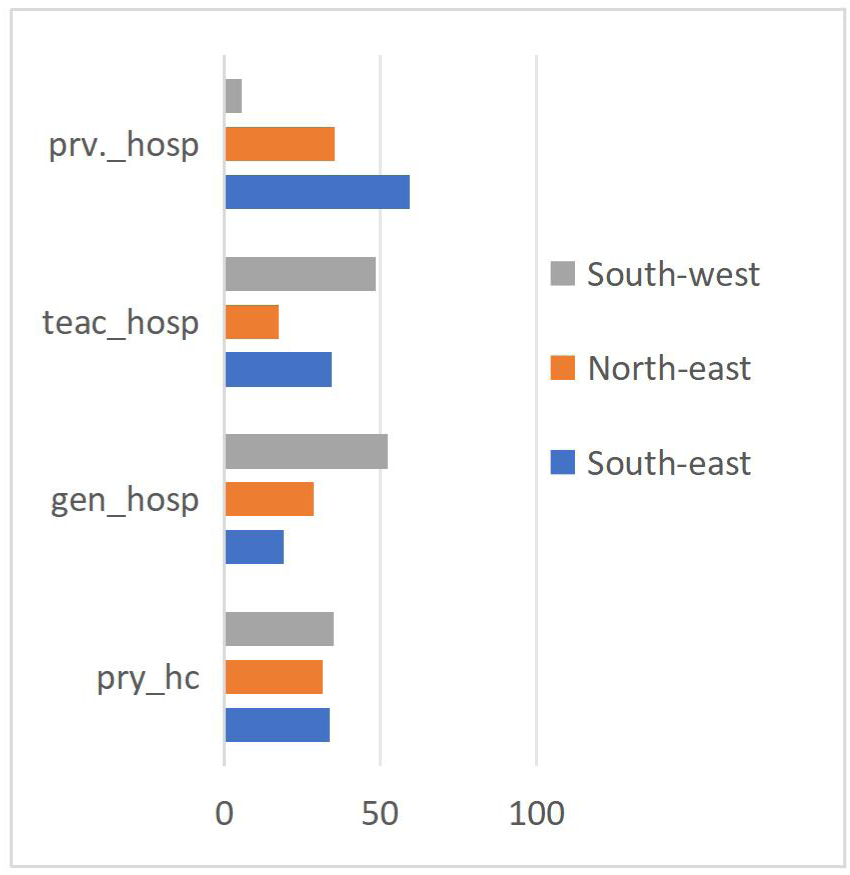
Access to healthcare by geopolitical zone. Note: pry_hc: primary healthcare; gen_hosp: general hospital; teac_hosp: teaching hospital; prv_hos:private hospital.

**Fig 6:**
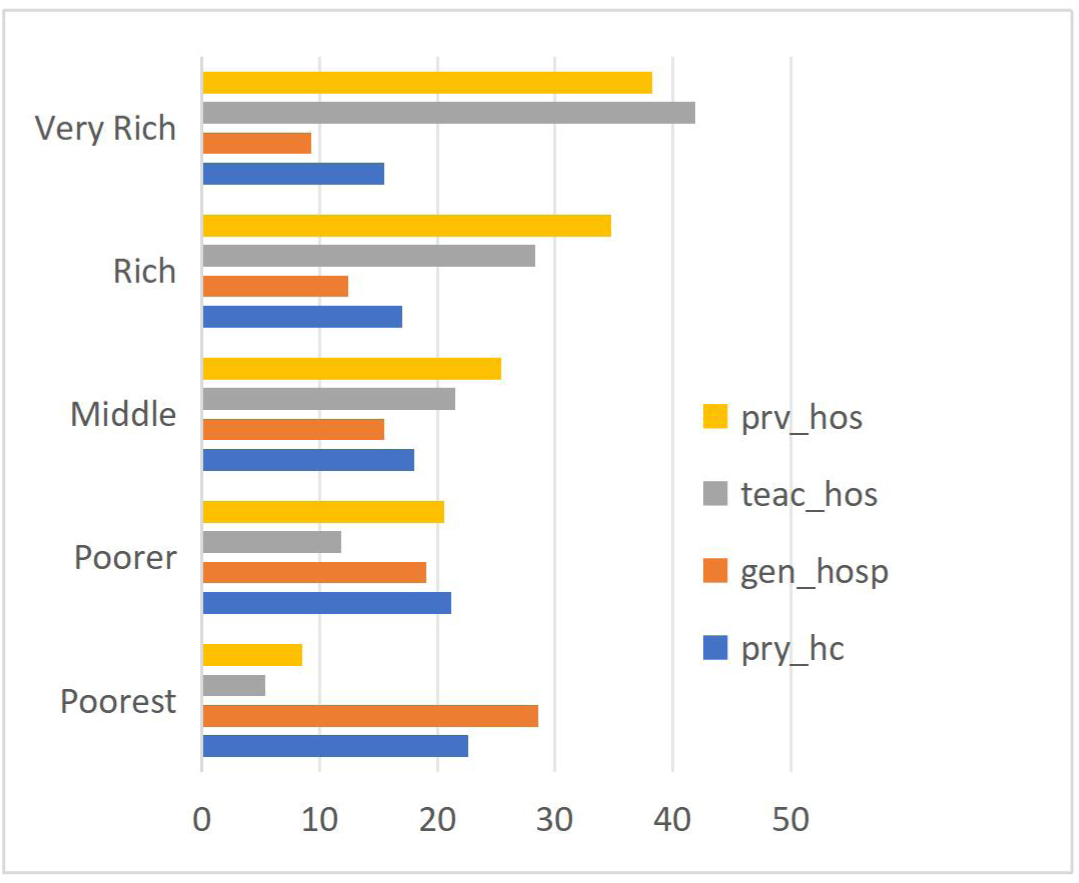
Access to healthcare by income quintile. Note: pry_hc: primary healthcare; gen_hosp: general hospital; teac_hosp: teaching hospital; prv_hos:private hospital.

**Figure 6a:**
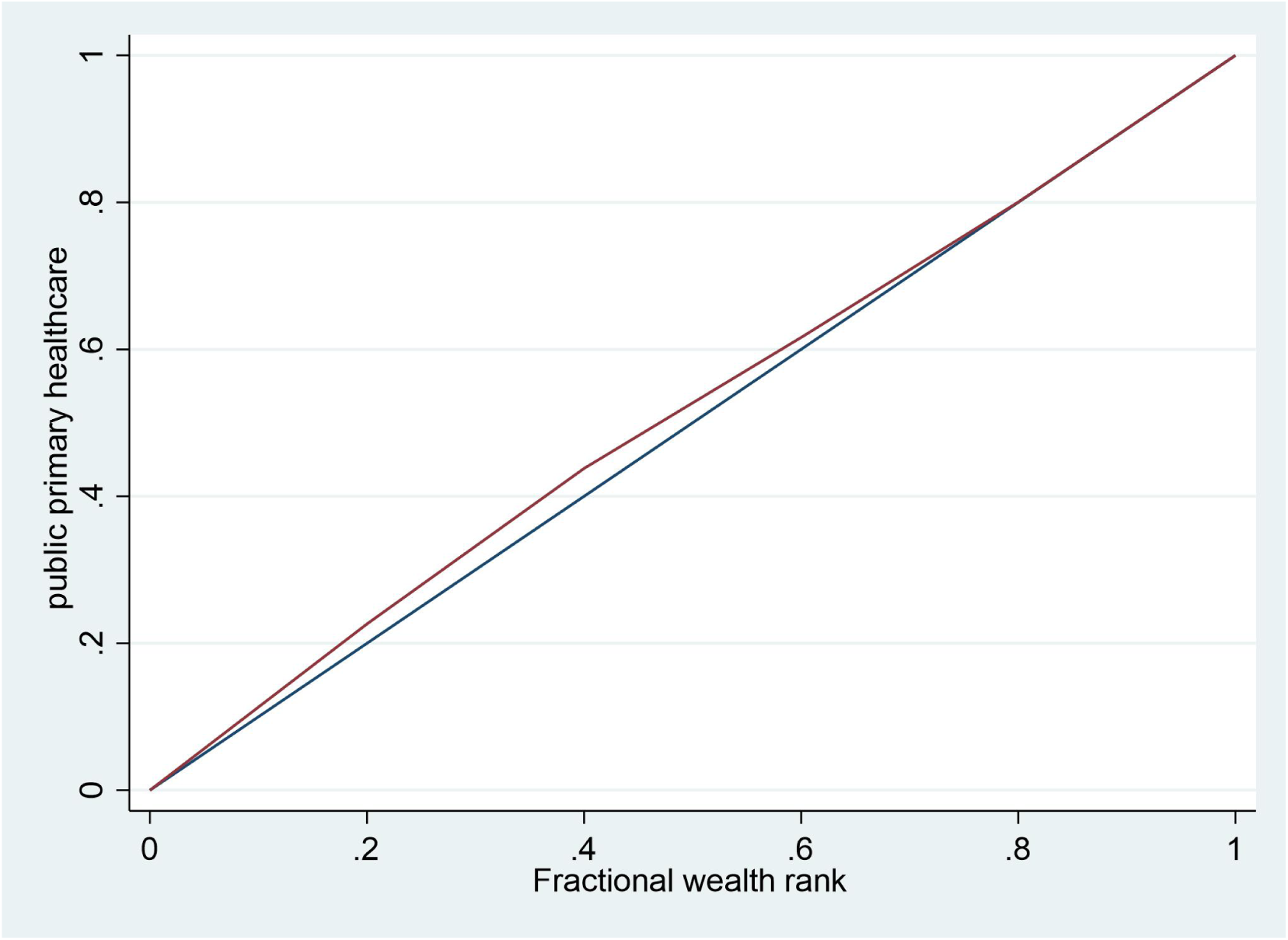
Concentration curve of public primary healthcare (dispensary/maternity)

**Figure 6b:**
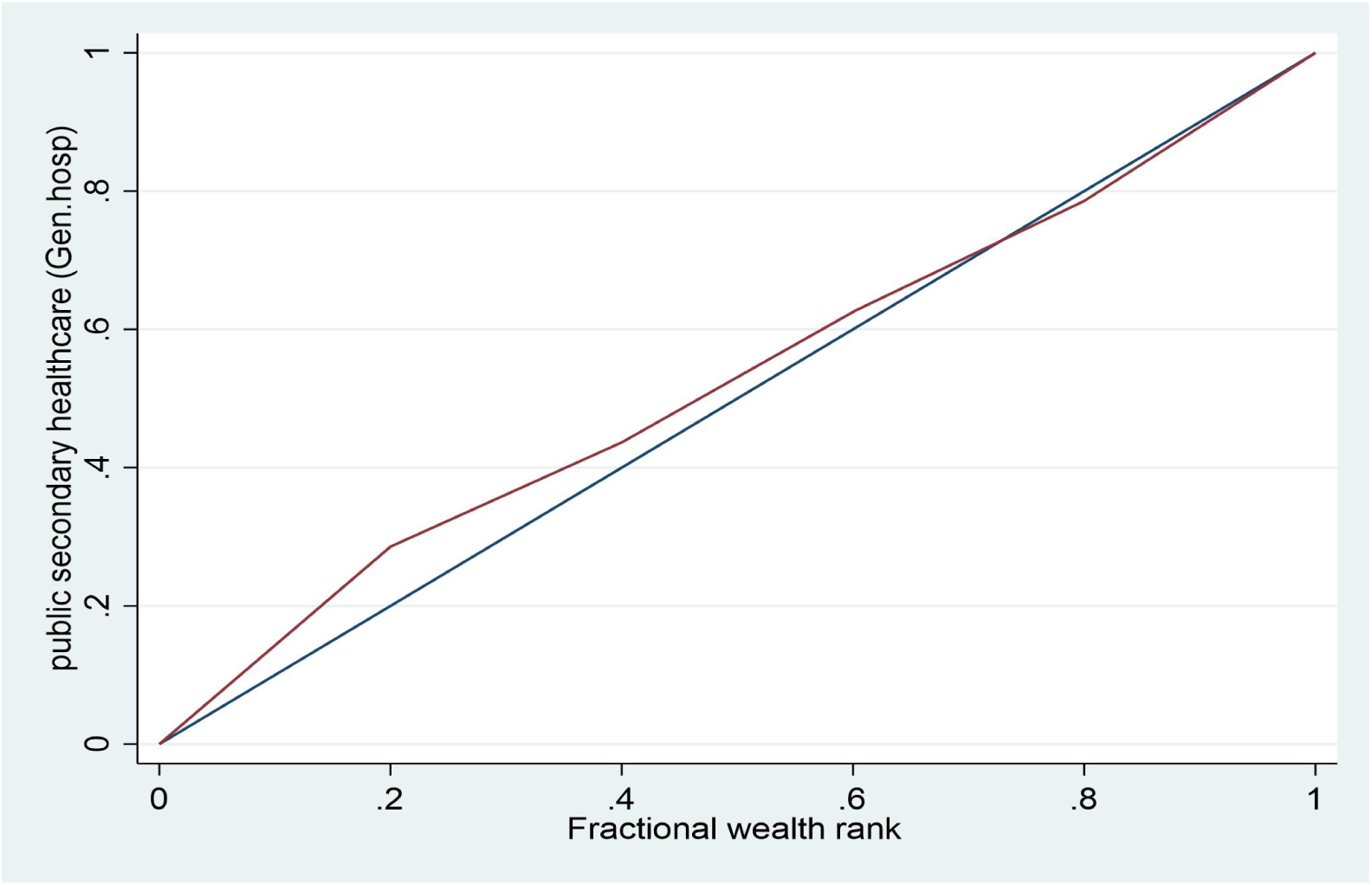
concentration curve of public secondary healthcare (general Hospital)

**Figure 6c:**
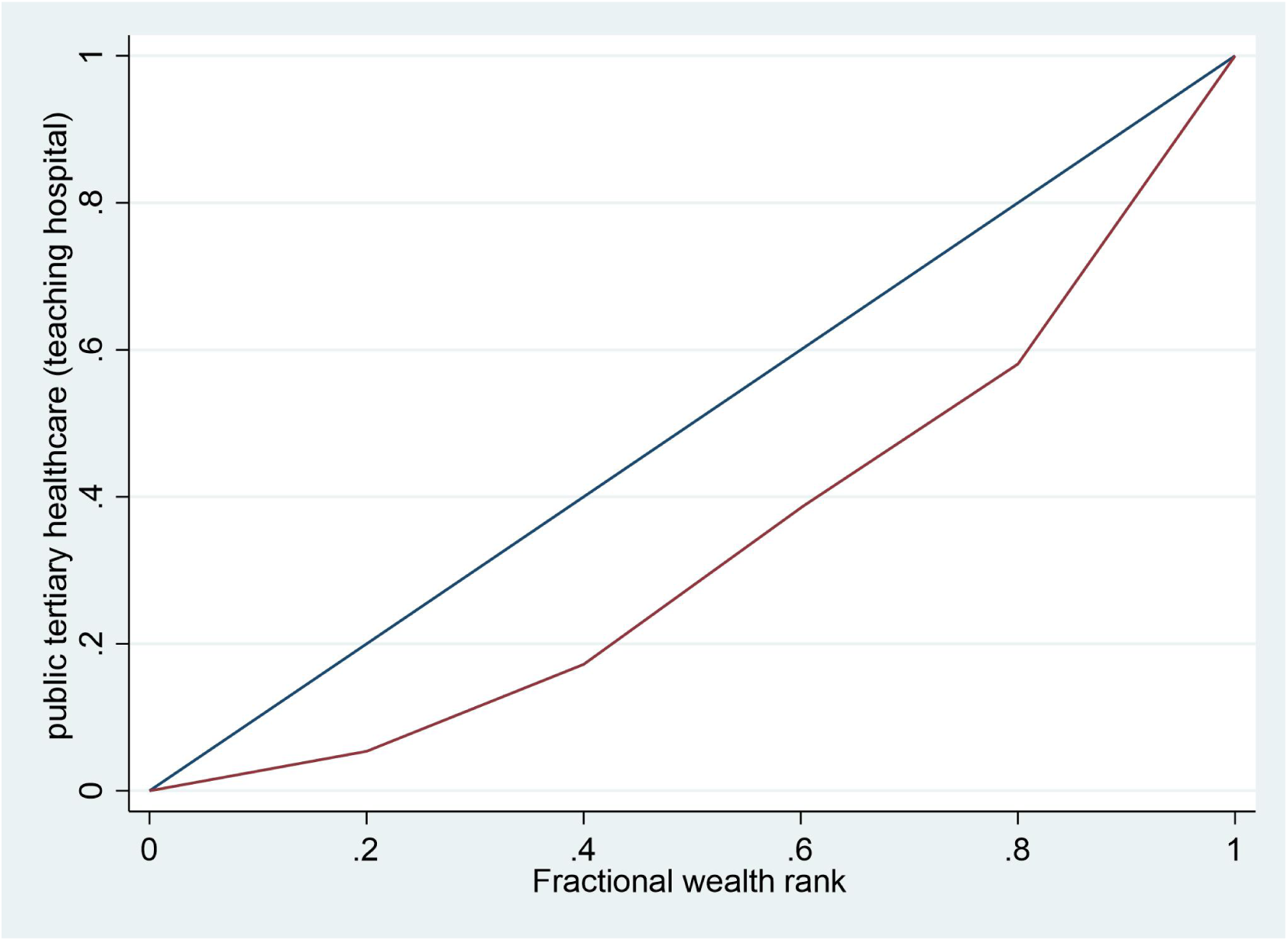
concentration curve of public tertiary healthcare (teaching hospital)

**Figure 6d:**
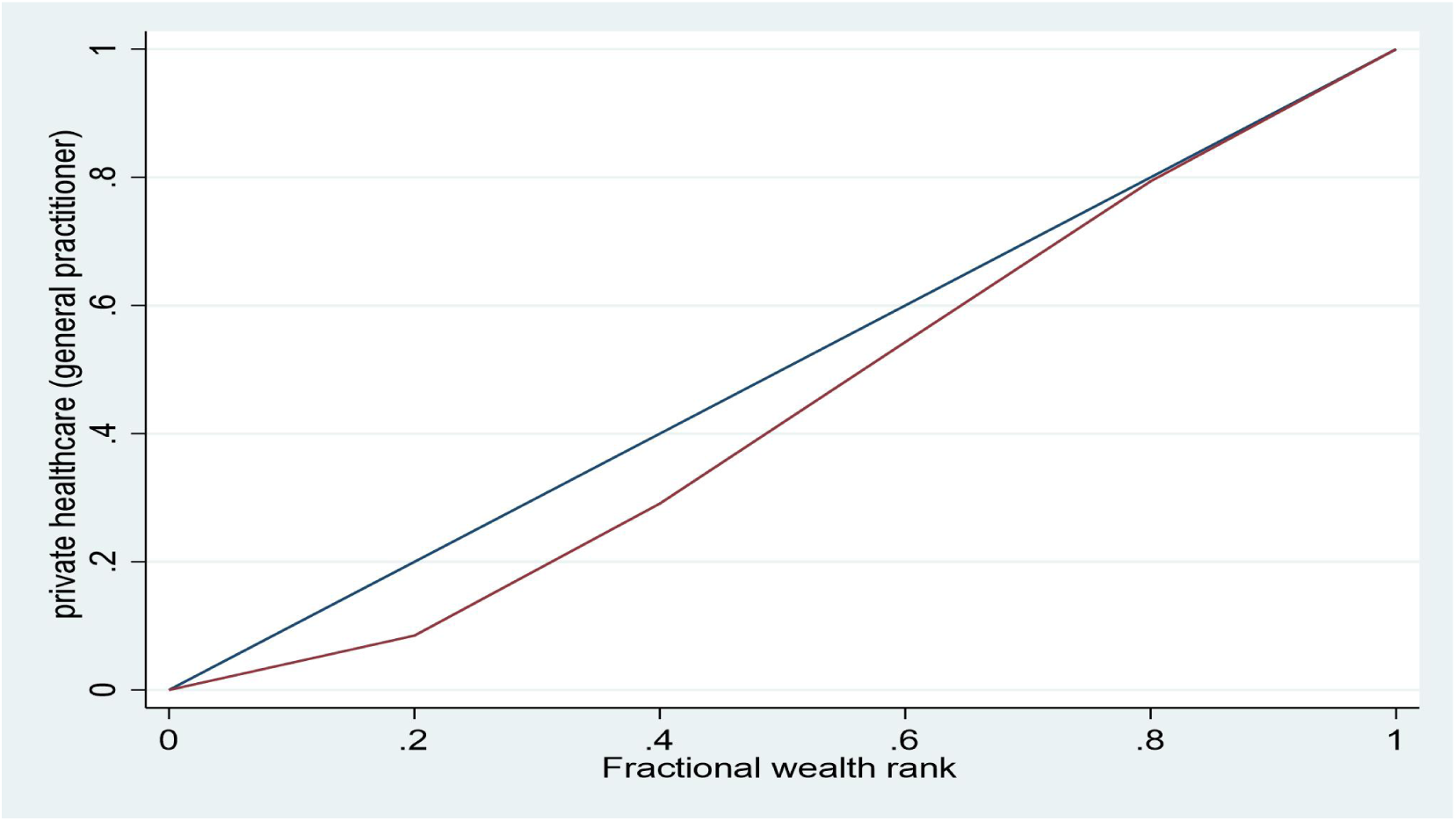
concentration curve of private general healthcare.

### 3.2 Inequality and Horizontal Inequity in access to healthcare

Concentration Index (CI) for primary (CI=-0.1054) and secondary healthcare (–0.1054) were negative and significant indicating pro-poor access. However, for tertiary (CI = 0.2382) and private general healthcare (CI = 0.1502), the CI were positive indicating a pro-rich access. The concentration curves in Figure 6a-6d corroborates what was indicated by the CI.

The greatest pro-rich inequality was observed for service at tertiary health facility (CI= 0.2382), while the greatest pro-poor inequality was observed for services at secondary (CI=-0.1063) health facility. When adjusted for differences in healthcare need, the HIs were less negative than CI for both primary (HI= – 0.0374) and secondary (HI= – 0.0377) healthcare implying less-pronounced pro-poor inequalities. This suggests diminishing inequality gap that tends towards fair distribution. For public tertiary (HI= 0.366) and private general healthcare (HI: 0.218), the HIs were more positive than CI indicating that for a given health need, access is more pro-rich widening the inequality gap in both care settings.

### 3.3 Decomposition of inequalities in access to inpatient healthcare

Table 3 shows the result of CI decomposition analysis. The first column shows the partial effect of socio-economic factor on access to various types of healthcare, the second column shows the concentration Index (CI) associated with each factor, the third column shows the absolute contribution of each factor to the overall CI and the fourth column shows the percentage contribution.

**Table 2:**
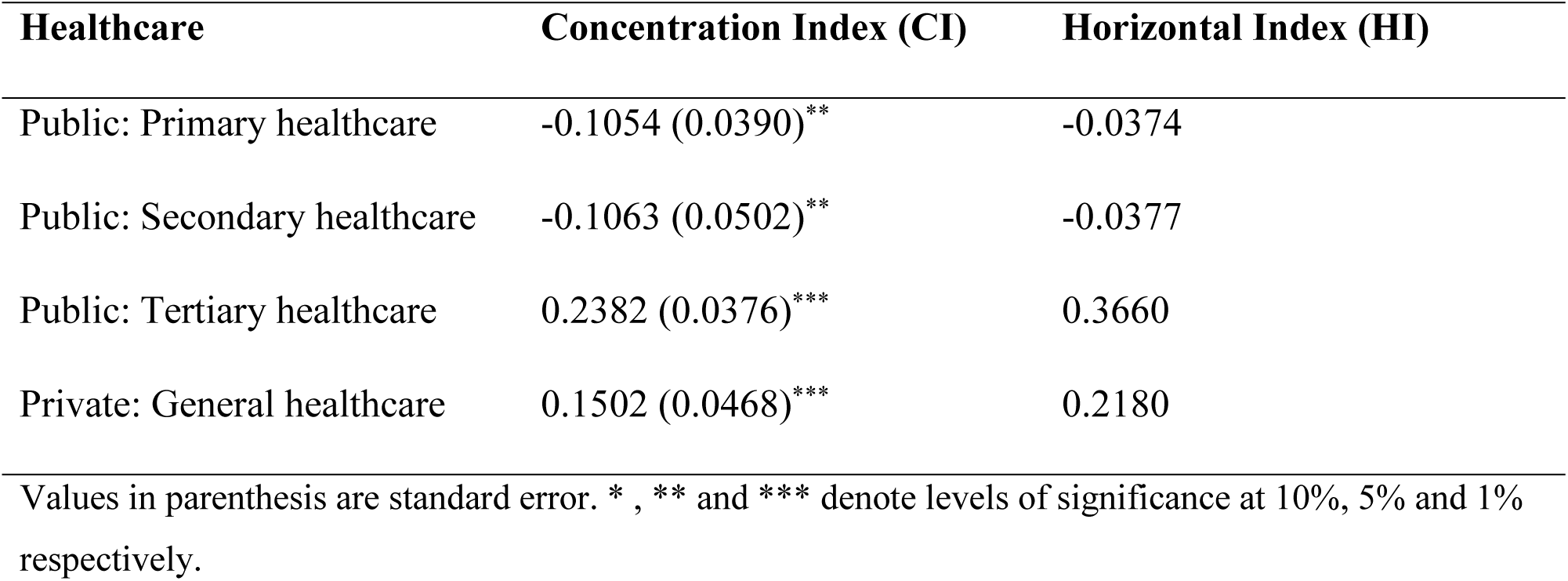
Unstandardized Concentration Index and Horizontal inequity Index.

| Healthcare | Concentration Index (CI) | Horizontal Index (HI) |
| --- | --- | --- |
| Public: Primary healthcare | -0.1054 (0.0390)** | -0.0374 |
| Public: Secondary healthcare | -0.1063 (0.0502)** | -0.0377 |
| Public: Tertiary healthcare | 0.2382 (0.0376)*** | 0.3660 |
| Private: General healthcare | 0.1502 (0.0468)*** | 0.2180 |

Values in parenthesis are standard error. \*, \*\* and \*\*\* denote levels of significance at 10%, 5% and 1% respectively.

**Table 3:**
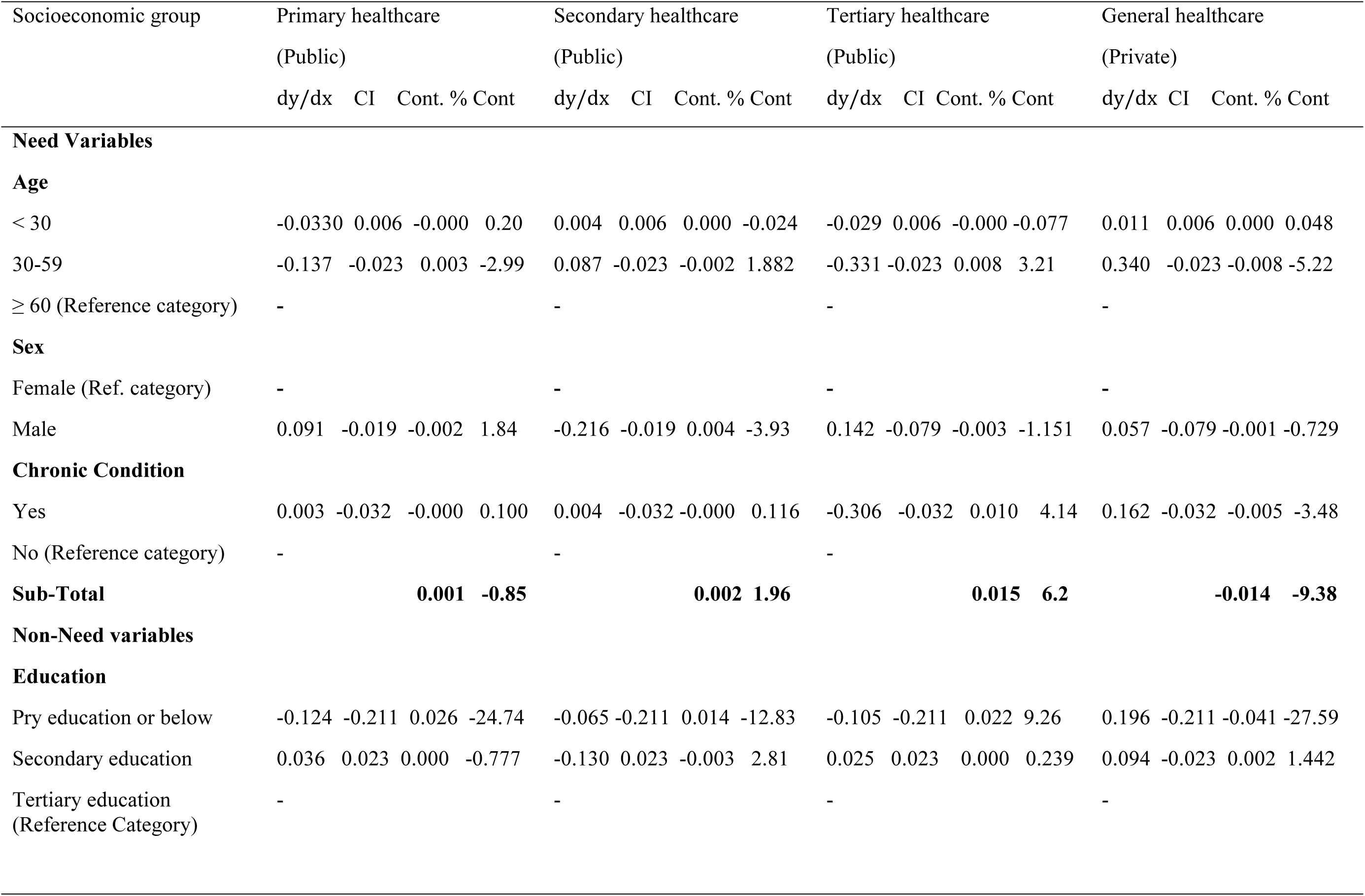
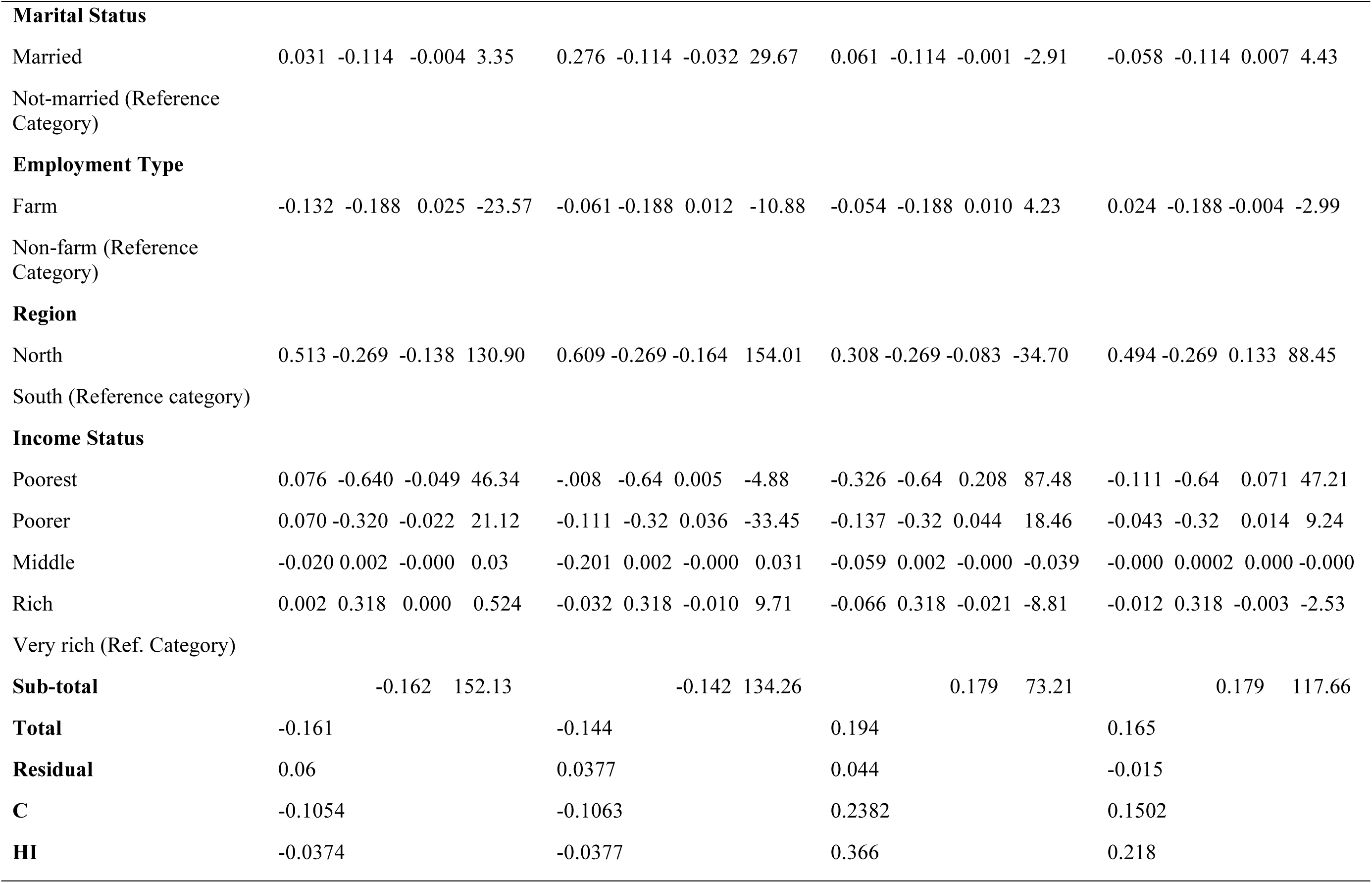
Decomposition of income-related inequalities in access to various types of healthcare in rural Nigeria.

Significant positive association was found between access to primary healthcare and age, educational level, employment type and region of residence. Older respondents (aged 60 and above), who had at least secondary education, primarily engaged in farming and resident in the northern region were more likely to access and use primary healthcare. Being male, married, middle-aged (30-59 years), attainment of primary education or below, engaged primarily in farming and reported chronic health condition and resident in the northern region were all concentrated among the poor. The need factor contributes marginally to pro-poor inequality [0.001(0.85%)], while the substantial proportion was explained by the non-need factor [-0.162 (152.13%)]. Of all the non-need factors, household economic status contributes the largest to pro-poor inequality. This is because better-off households were less likely to visit primary healthcare conditional on need making health resources more available to cater for the poorer patients.

Educational level, geographical region and economic status were significantly associated with secondary healthcare. Respondents who completed tertiary education, and those in the fifth quintile (very rich) of the income distribution were more likely to access the secondary healthcare. Being male, married, middle-aged (30-59 years), chronic health condition, primary education or below, primary engagement in farming and resident in the north were all concentrated among the poor. The Need factors [0.002(1.9%)] contributes very small amount to to pro-rich inequality, while the non-need factors explained very large pro-poor contributions to inequality. Of all the non-need factors, being resident in the north (–0.269) has the largest pro-poor contribution to inequality in the secondary healthcare.

Age, health condition, educational level, geographical region and economic status were all significantly associated with access to the tertiary healthcare. Older respondents aged 60 years and above, who reported chronic health condition, completed at least secondary education, resident in the south and those in the fifth quintile (very rich) of income distribution were more likely to access the tertiary healthcare. Being male, married, middle-aged (30-59 years), chronic health condition, having at most primary education, primarily engaged in farming, resident in the north were all concentrated among the poor. The non-need factors explained very large contribution [0.179(73.2%)] to pro-rich inequality in access to the tertiary healthcare. The pro-poor contributions of other non-need factors counteracts the pro-rich contributions. Of all non-need factors, household’s economic status [0.231(97%)] accounts for the largest contribution. Further, education, geographical region and economic status were significantly associated with access to the private general healthcare. Respondents who had at most primary education, resident in the south and very rich (5^th^ quintile) were more likely to access the private general healthcare. Being male, married, middle-aged (30-59 years), reported chronic health condition, primarily engaged in farming and resident in the north were all concentrated among the poor. The Non-need factor accounted for the largest pro-rich contribution [0.179(117.6%)]. Of all the non-need factors, being resident in the north (88.45%) accounted for the largest contribution to inequality in access to the private general healthcare.

## 4. Discussion

This study employed the Concentration Index (CI) and Horizontal Inequity (HI) to analyse income-related inequalities and inequities in access to the various types of healthcare. The study decomposed the inequalities in order to identify the contributions of socio-economic factors to inequalities in access to various types of healthcare. Findings show that the primary healthcare [public] is the most accessed, while the least accessed is the tertiary healthcare [public]. Across the various types of healthcare, access is low among female compared to the male, lowest among respondents who were below 30 years and among those with at most primary education. Overall, southwest had the most access to all types of public healthcare, southeast for private healthcare only, while the least access to any type of care was found among respondents resided in the northeast. We also found the poor households [those in the first and second quintile] dominated the primary and secondary healthcare,, while the rich households [those at the upper quintile of income distribution] dominated the tertiary [public] and private general healthcare. This result is similar to that reported by Yaya *et al.*[43] for Malawi and Ogundele *et al.* [44] for Nigeria and Ghana. Evidence of pro-poor inequalities was found in access to the public primary and secondary healthcare. When adjusted for the differences in health care need, the inequalities were less pro-poor and close to zero suggesting a fair distribution of health resources among different income groups. Our results are similar to that obtained by Bose and Dutta [45] for India; Zhang *et al.* [46] for China and Oburota *et al.* [32] for Nigeria but contrary to Guo *et al.* [47] for rural China. This could be attributed to the the impact of the recent reforms and efforts in the health sector allowing Nigeria to make notable progress towards achieving the Universal Healthcare Coverage (UHC).. These include the introduction of mandatory health insurance-National Health Insurance Authority-, Service Coverage Index which increased from 25% in 2003 to 44% in 2019; the 2018 Nigeria Health Workforce registry and increased fund for Basic Health Care Provisional Fund (BHCPF).

Further, the results also suggest that the wealthy and socio-economically advantaged people were more likely to seek care from private hospitals, where they are guaranteed of quality health services given their abilities to afford the user-fee leaving more resources for the poor and socio-economically disadvantaged people. However, evidence of significant pro-rich inequalities was found in access to public tertiary and private general healthcare. When adjusted for the differences in healthcare need, the inequalities were more pronounced (more pro-rich) suggesting that the rich people [upper quintile] were more advantaged compared to the poor [lower quintile]. These results are similar to that obtained by Johar *et al,* [48] for Indonessia; Li *et al.* [49] for rural China and Sharma *et al.* [50] for rura Bhutan.

While the result for the former set of healthcare shows considerable progress towards the attainment of the universal healthcare coverage (UHC), that of the latter suggests the need for special intervention in access to the tertiary healthcare in order to stem the growing trend of Non-Communicable Diseases (NCDs), which is projected to overtake infectious diseases by 2030 in Africa (Marquez and Farrington, 2013). Odunyemi *et al.* [51] also noted that Nigeria is witnessing a rising burden of NCDs with approximately one out of four Nigeria’s 30 to 69-years-olds risk premature death from Cardiovascular diseases (CVDs), cancers, respiratory diseases and diabetes.

Relatively older people were more likely to access and use primary and tertiary health facilities, while people with chronic health condition were less likely to access tertiary healthcare conditional on need. It is plausible to find the relatively older people making more use of primary healthcare given the increasing health need as they age [52]. People with primary education or below were less likely to access any type of public healthcare but more likely to access private healthcare conditional on need. These findings underscore the important role that education plays in people’s socio-economic gradient and the associated benefits including access to publicly-funded health facilities. It also highlights the need for renewed efforts by relevant authorities in achieving the universal basic education particularly in the rural area. These results are similar to that obtained by Yaya *et al.* [43] for Malawi, Ezzatabadi *et al.* [53] for Isfahan province in Iran and Nyamande *et al*. [54] in Northern Sweden. Turner *et al*. [55] also show that patients from educationally deprived areas face longer waits and receive less complex Emergency Department (ED) care.

Being resident in the southern Nigeria is associated with lower use of primary healthcare. It is not surprising to find the use of public primary healthcare unattractive for people resident in the southern Nigeria as majority are relatively wealthier than their northern counterparts and may prefer to use the service of private [general] or teaching hospital [tertiary], where access to improved health services including advanced clinical procedure and screening depends on the ability to pay out-of-pocket (OOP) healthcare expenses. Studies have also shown that deprived individuals are more likely to visit primary care conditional on need but much less likely to use specialist care than their wealthier counterparts [56, 57]. These findings are consistent with Mostafavi *et al.* [58] for Iran but contrary to Xu *et al.* [59] for rural China.

Through the decomposition analysis, we found the contribution of the need factor to be very insignificant across the the various types of healthcare, that of the non-need factor was overwhelming suggesting that, the disparities in access to healthcare were driven largely by factors outside the medical need. While economic status was found to make the largest pro-rich contribution, the region of residence has the largest pro-poor contribution to inequalities. The pro-poor contribution of the region of residence could be attributed to the concentration of the poor in the northern region. These findings are similar to that obtained by Abouie *et al.* [60] and Fu *et al.* [61] on inequalities in access to inpatient health services in China. For privately-funded genera health, the biggest source of pro-rich inequalities was economic status, while the biggest source of pro-poor inequalities was employment type. The pro-poor contribution of the employment type may be the result of concentration of the poor among the people primarily-engaged in farming occupation. These findings are consistent with IIinca *et al.,* [62].

## 5. Limitation of the Study

We acknowledges some limitations that are likely to shape the literature on health inequalities. First, while the study examined income-related inequalities and inequities in access to inpatient healthcare among households in rural Nigeria, the findings were based on data of access to inpatient care, making it difficult to upscale the outcome for the entire population that are normally affected by both the inpatient and outpatient care. Further, we used reported data for health condition of the respondents. It may be noted that reported data are subjective as they are based on self-perception, which may be affected by recall and measurement bias. Lastly, the data were collected at the height of insecurity situation in Nigeria making many communities and villages areas unsafe and inaccessible for survey data collection. As a result, the representative sample for the study could not be quarantined. Nonetheless, the sample size was appropriately weighted to account for sampling design. More over, since the political ideology and economic characteristics of the country are still patterned along the north-south divide, the country can broadly be divided into North and South. Despite the limitations, the study provides useful insights necessary to shape the policy making decision for closing the inequality gap in healthcare.

## 6. Conclusion and implications

The study found evidence of pro-poor inequalities in access to inpatient public primary and secondary healthcare and more pronounced when adjusted for differences in healthcare need. For access to public tertiary and private general healthcare, the inequalities are pro-rich, and more pronounced when adjusted for differences in healthcare need. The inequalities in access to the various types of healthcare were driven mainly by the non-need factors. Economic status and region of residence were the largest sources contributing to pro-rich and pro-poor inequalities respectively. The foregoing suggests the need to prioritize health interventions in favour of the people living in the northern part of the country and in particular the rural areas. It also suggests the need to scale up and broaden the scope of health insurance coverage for healthcare at the tertiary level.

## Data Availability

All data produced in the present study are available upon reasonable request to the authors.

